# Interventions to improve men’s engagement in HIV and other key services in sub-Saharan Africa: a scoping review

**DOI:** 10.1101/2023.10.25.23297534

**Authors:** Kathryn Dovel, Julie Hubbard, Lycias Zembe, Nathan Ford, Morna Cornell, Will Belshe, Lawrence Long, Stephanie Davis, Paula A. Munderi, Rachel Baggaley, Wole Ameyan

## Abstract

**Background:** Men in sub-Saharan Africa (SSA) continue to have worse health outcomes across HIV, STI, and TB-HIV co-infections as compared to women. Improving service coverage is critical for population health and HIV epidemic control. In HIV, for example, recent models show that improving men’s HIV testing and treatment coverage could reduce HIV incidence among women in the region by half. There is potential to combine and optimize services across HIV, STI and TB-HIV co-infections, yet little is known about effective interventions to improve men’s outcomes across health services.

**Methods:** We conducted a scoping review of interventions to understand what interventions work for men, and any synergies in interventions that work across health services. We specifically focused on interventions aimed to improve service utilization in the following service domains: condom use; pre-exposure prophylaxis (PrEP); STI testing and treatment; HIV testing, initiation, and retention; and TB testing and treatment among those living with HIV (co-infected). Articles and abstracts had to include sex-disaggregated data or solely focus on men’s health service outcomes. We searched PubMed, Medline, Cochrane Central Register of Controlled Trials, the CABI Global Health databases, and major international conference abstracts. We included studies from SSA, published between January 1, 2009 to Dec 31, 2022, quantitative data on at least one of the selected service domains, disaggregated data for the general male population (not solely key populations), an intervention study (report outcomes for at least one non-standard service delivery strategy) with a comparison group, and available in English. We describe the type of interventions evaluated and synthesize overarching themes of “what works” for reaching men.

**Findings:** We identified 15,595 intervention articles and included 71 in the scoping review, representing 111 unique interventions. Over a quarter of interventions targeted male partners and only 7 exclusively targeted men. Nearly half of the interventions had HIV testing as their primary outcome. Only a handful of interventions included outcomes related to condom use, STI, or TB co-infection services. No interventions examined the effect of PrEP use among general male populations. Community services was the most common intervention type (n=40, 36%), followed by community outreach (n=19; 17%), incentives (n=16; 14%) and facility services (n=16, 14%). Counseling and peer support had the least number of interventions evaluated (n=8, 7%). We were unable to identify cross-cutting strategies to reach men across HIV and related health services in sub-Saharan Africa, largely because there is little evidence outside HIV testing interventions. The limited evidence available points to the fact that men need convenient, active outreach, and improved experiences with health services. The same principles may apply to all services intended to reach men, including sexual health and TB co-infection services, although the evidence is limited.

**Conclusion:** This review highlights the need for additional research on cross-cutting strategies to improve men’s engagement in HIV and related health services. The limited evidence available suggests that convenient services, actively engaging men, and providing positive experiences with health services largely improve service utilization. Additional evidence is needed for PrEP use and non-HIV services (such as STI and TB co-infection).

## Introduction

Men in sub-Saharan Africa (SSA) experience higher rates of morbidity and mortality from HIV, TB, and higher rates of many sexually transmitted infections (STIs).^1–6^ as compared to women. Among those infected with HIV, men are twice as likely as women to have advanced HIV disease^7^ and twice as likely to die from TB.^8,9^ Untreated STIs among men are a major contributor to HIV acquisition.^10^ Addressing disparities in HIV and related services is critical for population health and HIV epidemic control. Recent models show that improving men’s HIV testing and treatment coverage could reduce HIV incidence among women in the region by half.^11^

An important next step is to identify and implement evidence-based strategies to improve men’s use of health services across the HIV cascade and related health services, such as STI and TB-HIV co-infection. There is extensive synthesis of what works for men’s use of HIV testing services^12–16^ and growing evidence around linkage^17–19^ and retention^20^ to HIV treatment services. There is some evidence about strategies that work to reduce sexual risk behavior among adolescent and young men.^21^ However to our knowledge there is little synthesized evidence around what works for HIV-related services such as TB co-infection and STI services, and little to no synthesis of what works across the HIV cascade and related health services.

Identifying cross-cutting strategies that work for men across the HIV cascade and related health services is important. It can promote scalability and sustainability, moving away from siloed interventions and moving toward holistic health systems approaches. Fragmented health systems reduce the ability of health services to respond to the needs of vulnerable clients.^22^ Sustainable development goals acknowledge that improving health means improving multiple components of health and health outcomes. Further, men’s experiences with health services are not siloed. Positive or negative experiences with any health service can impact men’s decision to use HIV services in the future.^23–25^ Improved experiences with general health services (not just HIV services) may improve men’s engagement with health systems more broadly, and HIV services specifically.

We conducted a scoping review of literature from 2009 to 2022 to examine the impact of interventions on men’s use of HIV and other key services (i.e., condom use and PrEP services; STI testing and treatment; HIV testing, initiation, and retention; and TB-HIV co-infection testing and treatment) in sub-Saharan Africa. We identify potential synergies and optimized delivery strategies across HIV and other key services and identify key research gaps to facilitate cross-cutting interventions for men.

## Methods

We used the extended framework by Levac and colleagues^26,27^ in accordance with the Preferred Reporting Items for Systematic Reviews and Meta-analysis extension for scoping reviews (PRISMA-ScR) checklist.^28^ This review is drawn from larger work to identify interventions that facilitate the use of HIV and related services among men in sub-Saharan Africa conducted to support the development of the World Health Organization’s 2023 Men and HIV Framework. Here we describe findings from the scoping review focused on HIV and related services. We focus on articles that report intervention impact on men’s use of one or more HIV or related services in SSA. We exclude articles on voluntary medical male circumcision (VMMC) because, unlike other services, VMMC is only needed once across the life course and barriers/facilitators to service utilization are very different than other services.

### Search strategy and data selection

We included studies of interventions that included some comparison data. Most studies were randomized control trials, quasi-experimental studies, observational cohort studies, and program evaluations. All articles described interventions to improve the use of the following services in SSA: condom use; pre-exposure prophylaxis (PrEP); STI testing and treatment; HIV testing, initiation, and retention; and TB testing and treatment among those living with HIV (co-infected). All articles included at least one intervention (i.e., non-standard service delivery strategy) with a comparison group – comparisons were not required to be standard of care. We excluded articles that exclusively targeted key populations as these populations may need different interventions than the general male population. Articles may include both male and female participants but were only included if they reported sex-disaggregated results. Finally, we only included articles where participants were from SSA (using the United Nations sub-regions of Africa) and articles were available in English. We retrieved articles published between January 1, 2009 to Dec 31, 2022.

To identify articles, we searched the PubMed database and then adapted our search strategy for Medline, Cochrane Central Register of Controlled Trials, the CABI Global Health databases, and major international conference abstracts. We searched using Medical Subject Headings (MeSH), keywords, and filters as appropriate (see Table 1 for keywords). We completed two searches: We first searched for sources published from January 1, 2009, to the date of the literature pull (May 9, 2021). We then completed a revised search from the date of the last literature pull (May 9, 2021) until Dec 31, 2022.

**Table 1.** Key search words used.

| Concept | Text Key Words |
| --- | --- |
| ART | ART; Antiretroviral; Retroviral; HAART |
| HIV | HIV; human immunodeficiency virus |
| Condom use | Safe* sex; Condom; Pre-exposure prophylaxis; Sexual partner* |
| PrEP | Pre-exposure prophylaxis; PrEP |
| STI | Sexually transmitted infection* (Bacterial Vaginosis, Chlamydia, Gonorrhea, Hepatitis, Herpes, Mycoplasma genitalium, Syphilis, Trichomoniasis) |
| TB-HIV co-infection | Tuberculosis AND HIV; co-infection |
| Initiation | Initiat*; Uptake; Link*; Early Care |
| Retention | Retention; Retain*; Viral suppression; Sustained Virological Response"; advanced HIV; Acquired immunodeficiency syndrome |
| Sub-Saharan Africa | *Saharan Africa ( Angola, Benin, Botswana, Burkina Faso, Burundi, Cabo Verde, Cameroon, Central African Republic, Chad, Comoros, Congo, Democratic Republic of, Congo, Republic of Cote d'Ivoire, Equatorial Guinea, Eritrea, Eswatini (Formerly Known as Swaziland), Ethiopia, Gabon, Gambia, The, Ghana, Guinea, Guinea-Bissau, Kenya, Lesotho, Liberia, Madagascar, Malawi, Mali, Mauritania, Mauritius, Mozambique, Namibia, Niger, Nigeria, Rwanda, Sao Tome and Principe, Senegal, Seychelles, Sierra Leone, Somalia, South Africa, South Sudan, Sudan, Tanzania, Togo, Uganda, Zambia, Zimbabwe) |
| Intervention | Intervention*; Trial; Random*; Program*; Project; Strateg* |
| Publication year limit | 2009:2021.(sa_year); 2021; 2022 (sa_year) |

### Selection of sources of evidence

We first removed duplicates. One author then conducted a title and abstract review and removed sources that did not meet the study inclusion and exclusion criteria. We entered results from the search strategy into an Endnote^29^ database and two reviewers reviewed the full text of the remaining sources to determine final study eligibility, with any conflicts resolved by KD.

### Data charting process

We developed a data-charting form (Excel spreadsheet) based on the study objectives. Information included: study and participant characteristics (e.g., country, year of data collection, eligibility criteria), details of intervention and comparison models (e.g. location, frequency, and services provided), health service or clinical outcomes captured, and any concerns regarding quality (including potential bias in data collection, reporting, or analysis). Data were entered directly into the structured data-charting form independently by four reviewers. After all data were entered into the data-charting form, lead authors reviewed and refined the form iteratively. Any inconsistencies were reconciled and KD performed spot-checks.

### Synthesis

We categorized interventions into five intervention types, which were informed by Duncombe’s key components of tailored care across the treatment cascade^30^: facility-based services, community-based services, community outreach, counseling and/or peer support, mHealth, and incentives (see Table 2 for definitions). We summarized interventions, key components, and key limitations across service domains.

**Table 2.** Intervention type and definitions.

| Intervention Type | Definition |
| --- | --- |
| Facility-based services | Interventions that primarily focused on changes to protocols or services within facilities. |
| Community-based services | Biomedical services delivered outside the health facility. |
| Community outreach | Community-based activities to encourage use of biomedical services at health facilities, such as community sensitization campaigns, tracing activities, and facility referrals. |
| Counseling/peer support | <u>Ongoing</u> counseling and/or peer engagement to encourage positive health outcomes, such as building demand creation, health literacy, self-efficacy and agency, and identifying and developing plans to overcome barriers to care. |
| Incentives | At least one opportunity to receive monetary/non-monetary incentives conditional on one's use of HIV or related health services. |
| mHealth | Using mobile devices, tablets, and other technological devices to offer health services, counseling, monitoring and evaluation, or reminders, etc. |

## Results

We identified 15,595 intervention articles. We removed 4,406 duplicates, reviewed 11,187 titles, and 245 data sources qualified for full review. Under full review we excluded 174 data sources (Fig 1): not focused on men’s health (n=15), data not disaggregated by sex (n=87), non-intervention study (n=36), no comparison group (n=25), not in sub-Saharan Africa (n=7) and only included key populations (n=4) (see Figure 1).

**Figure 1.**
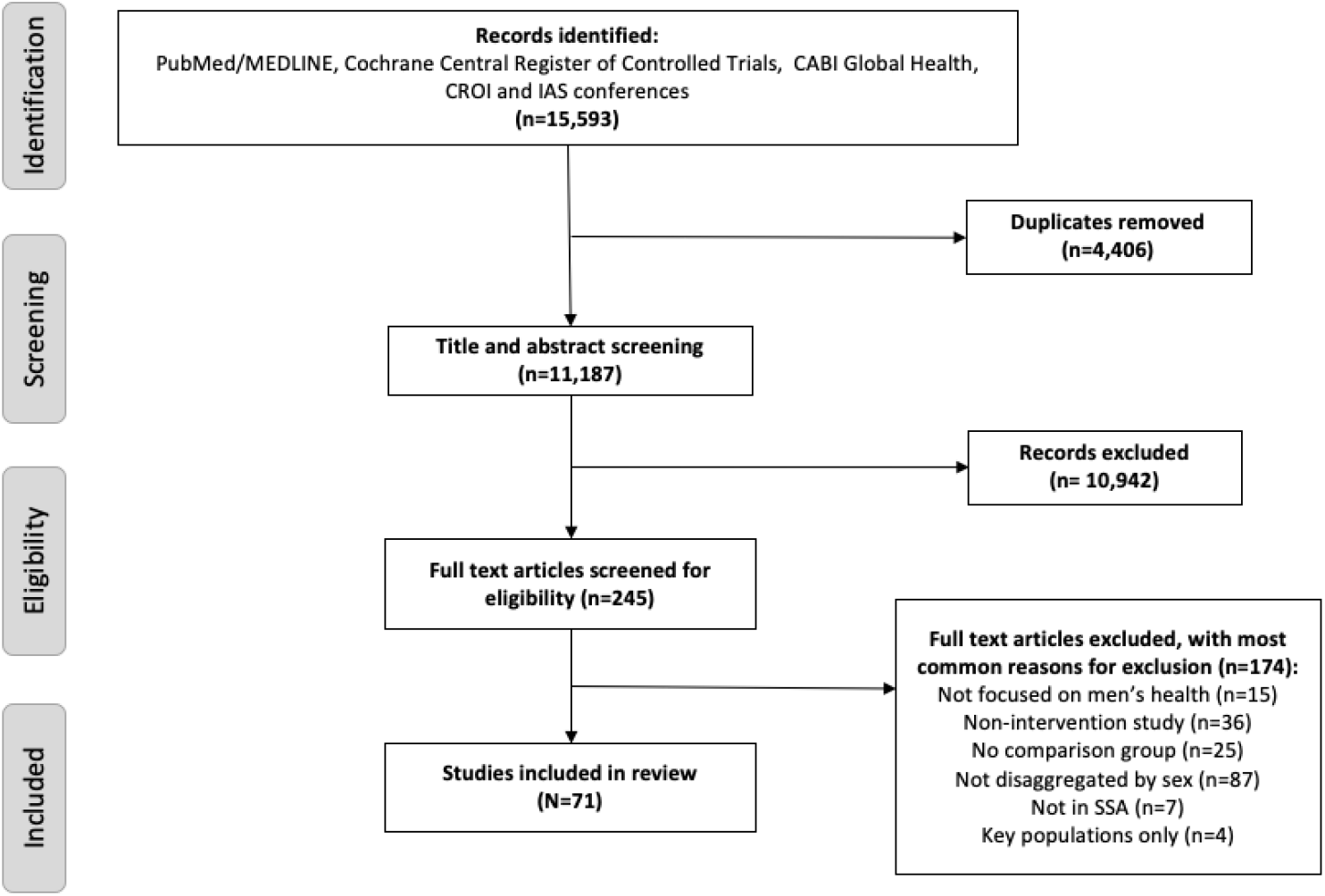
PRISMA flow diagram

### Settings and Populations of Included Interventions

A total of 71 articles were included in the review. Nearly all studies were conducted in eastern and southern Africa (n=70, 99%) and a large proportion of articles were published after 2016 (n=55, 77%), representing interventions within the era of universal treatment (see Table 3). Only 7 (10%) studies examined interventions tailored exclusively to men; these focused on HIV testing (n=7) and ART linkage (n=2). A total of 19 (27%) studies targeted male partners of pregnant and/or breastfeeding women or women living with HIV, 4 (6%) studies explicitly targeted adolescents and young men and no interventions explicitly targeted middle and older age men (not shown). Most studies focused on HIV services (n=64, 90%): among those, the majority had HIV testing services as the primary outcome (n=51, 80%). Only 2 studies focused on condom use, 2 on STIs, and 3 on TB-HIV co-infection. No interventions examined the impact on PrEP for men. About 62% (n=44) were randomized control trials.

**Table 3.** Description of settings and populations included in articles (n=71)

| Service Domain | Total | Published<br>>2016 | Male Only<br>Intervention | Eastern/Southern<br>Africa | West Africa | Central Africa |
| --- | --- | --- | --- | --- | --- | --- |
| Increase condom use | 2 | 0 | 0 | 2 | 0 | 0 |
| PrEP | 0 | 0 | 0 | 0 | 0 | 0 |
| STI | 2 | 0 | 0 | 2 | 0 | 0 |
| HIV/TB-co infection | 3 | 0 | 0 | 3 | 0 | 0 |
| HIV testing | 51 | 14 | 6 | 50 | 1 | 0 |
| HIV linkage to care | 10 | 1 | 2 | 10 | 0 | 0 |
| HIV retention and re-engagement | 3 | 0 | 0 | 3 | 0 | 0 |
| **Total** | **71** | **15** | **8** | **70** | **1** | **0** |

### Interventions Included

A total of 111 interventions were evaluated (see Table 4). Community services was the most common intervention type (n=40, 36%), followed by community outreach (n=19; 17%), incentives (n=16; 14%) and facility services (n=16, 14%). Counseling and peer support had the least number of interventions evaluated (n=8, 7%). Interventions varied by type of service type.

**Table 4.** Description of interventions by intervention type (n=111)

| **Service Domain** | **Total** | **Facility Services** | **Community Services** | **Community Outreach** | **Counseling/ Peer Support** | **Incentives** | **mHealth** |
| --- | --- | --- | --- | --- | --- | --- | --- |
| Increase condom use | 4 | 0 | 0 | 0 | 4 | 0 | 0 |
| PrEP | 0 | 0 | 0 | 0 | 0 | 0 | 0 |
| STI | 5 | 2 | 0 | 3 | 0 | 0 | 0 |
| TB-co infection | 3 | 1 | 1 | 0 | 0 | 0 | 1 |
| HIV testing | 82 | 7 | 37 | 16 | 3 | 12 | 7 |
| Linkage to care | 12 | 6 | 2 | 0 | 0 | 3 | 1 |
| Retention | 5 | 0 | 0 | 0 | 1 | 1 | 3 |
| **Total** | **111** | **16** | **40** | **19** | **8** | **16** | **12** |

Characteristics of included studies and study results are summarized in Table 5 and 6.

**Table 5.**
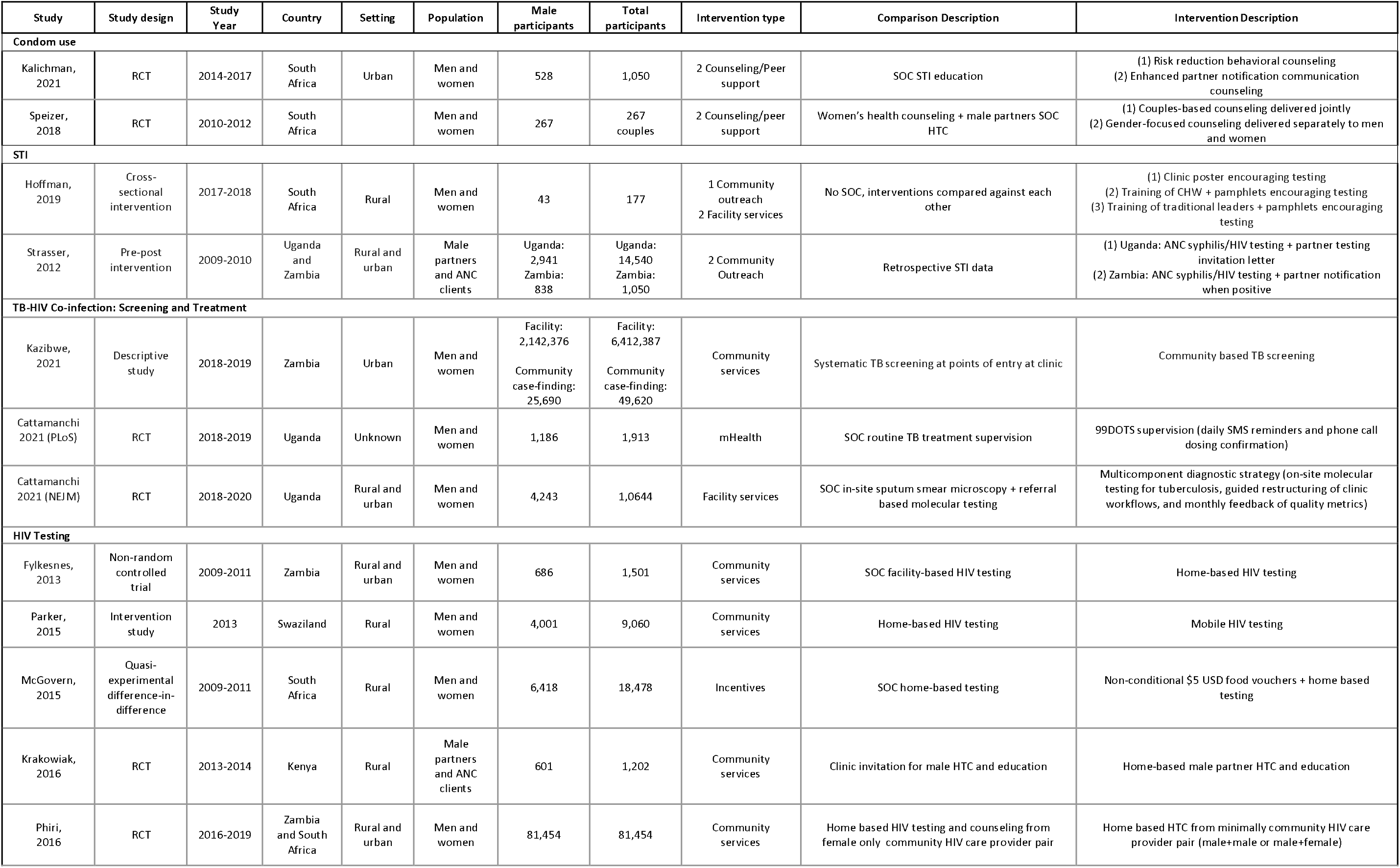
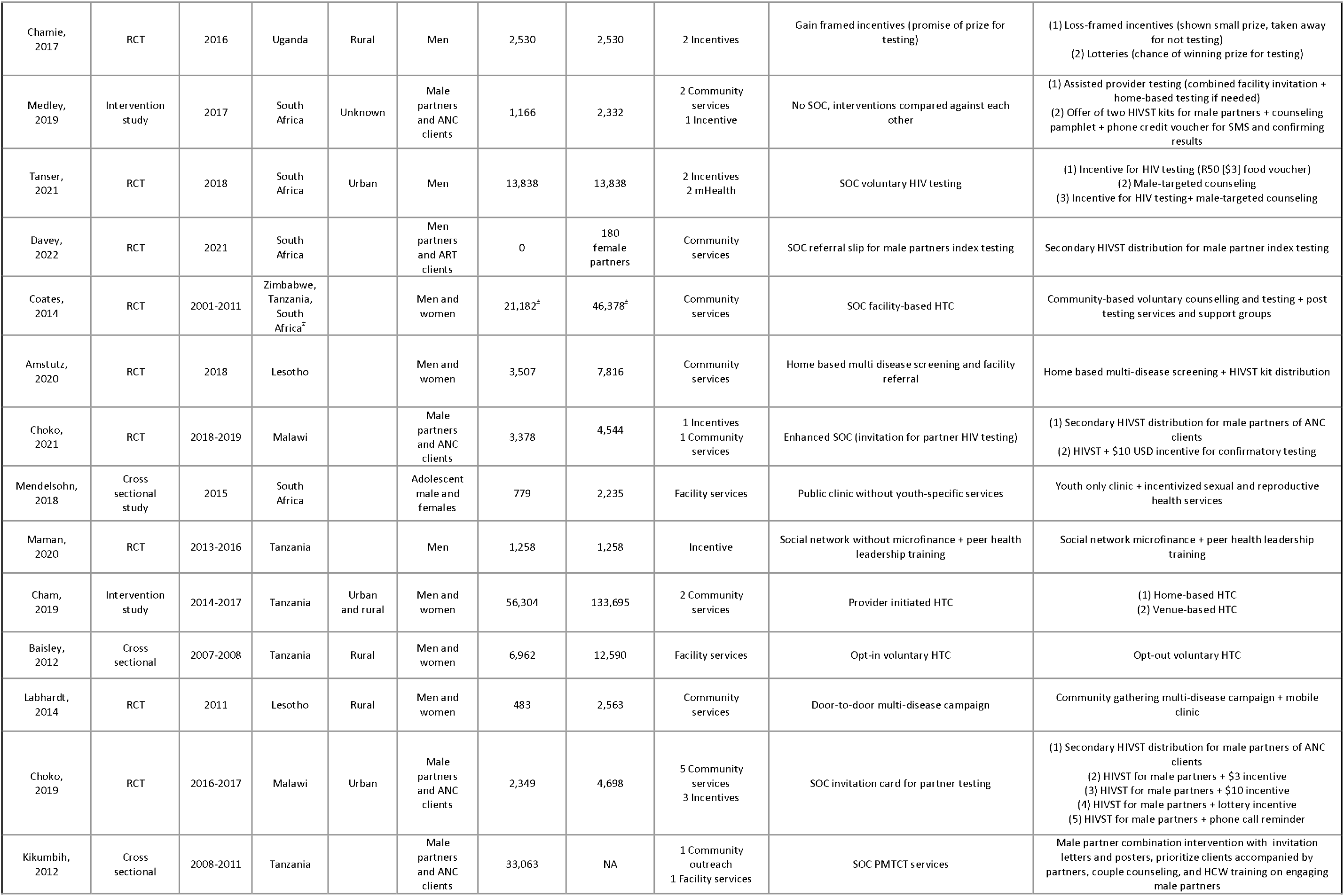
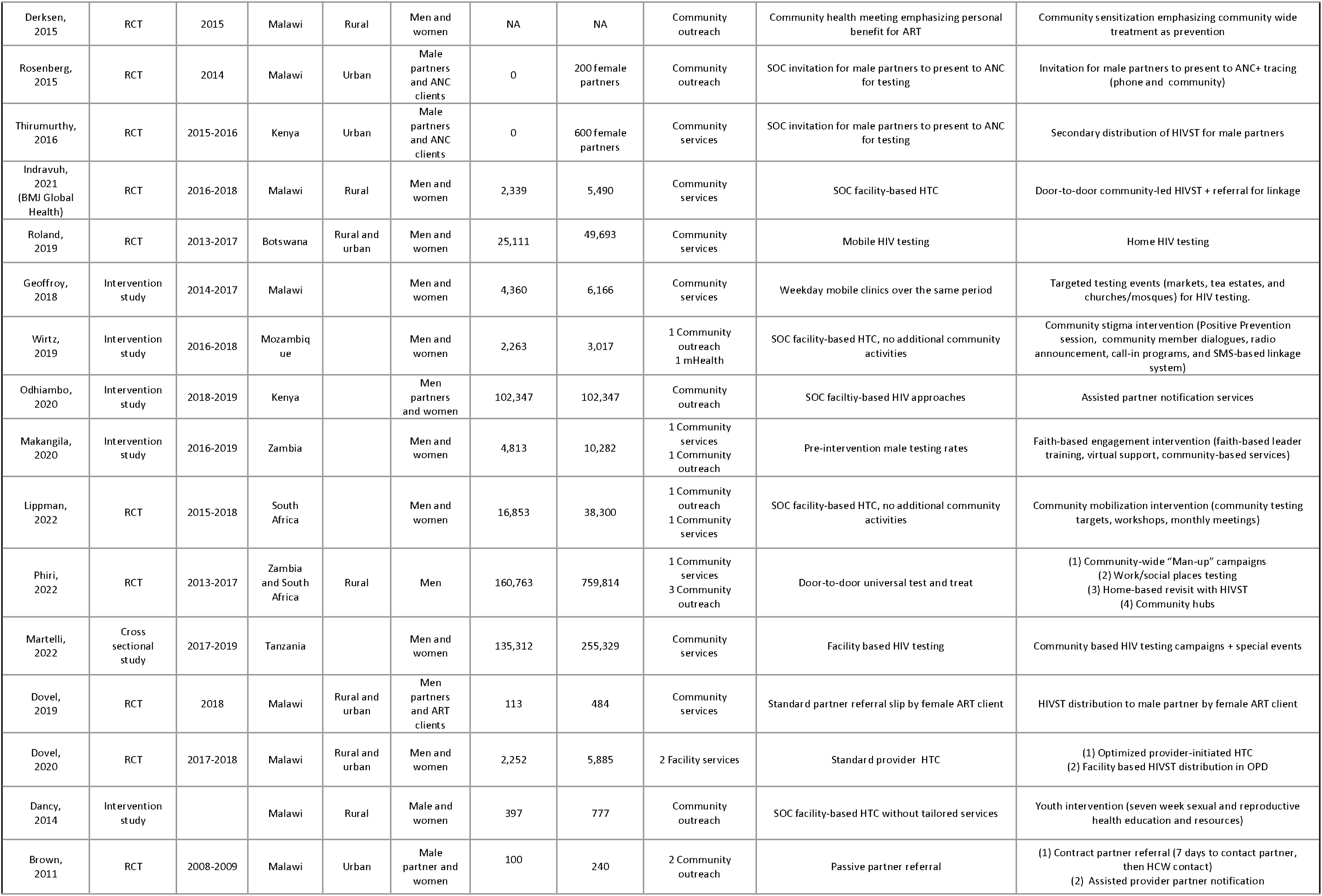
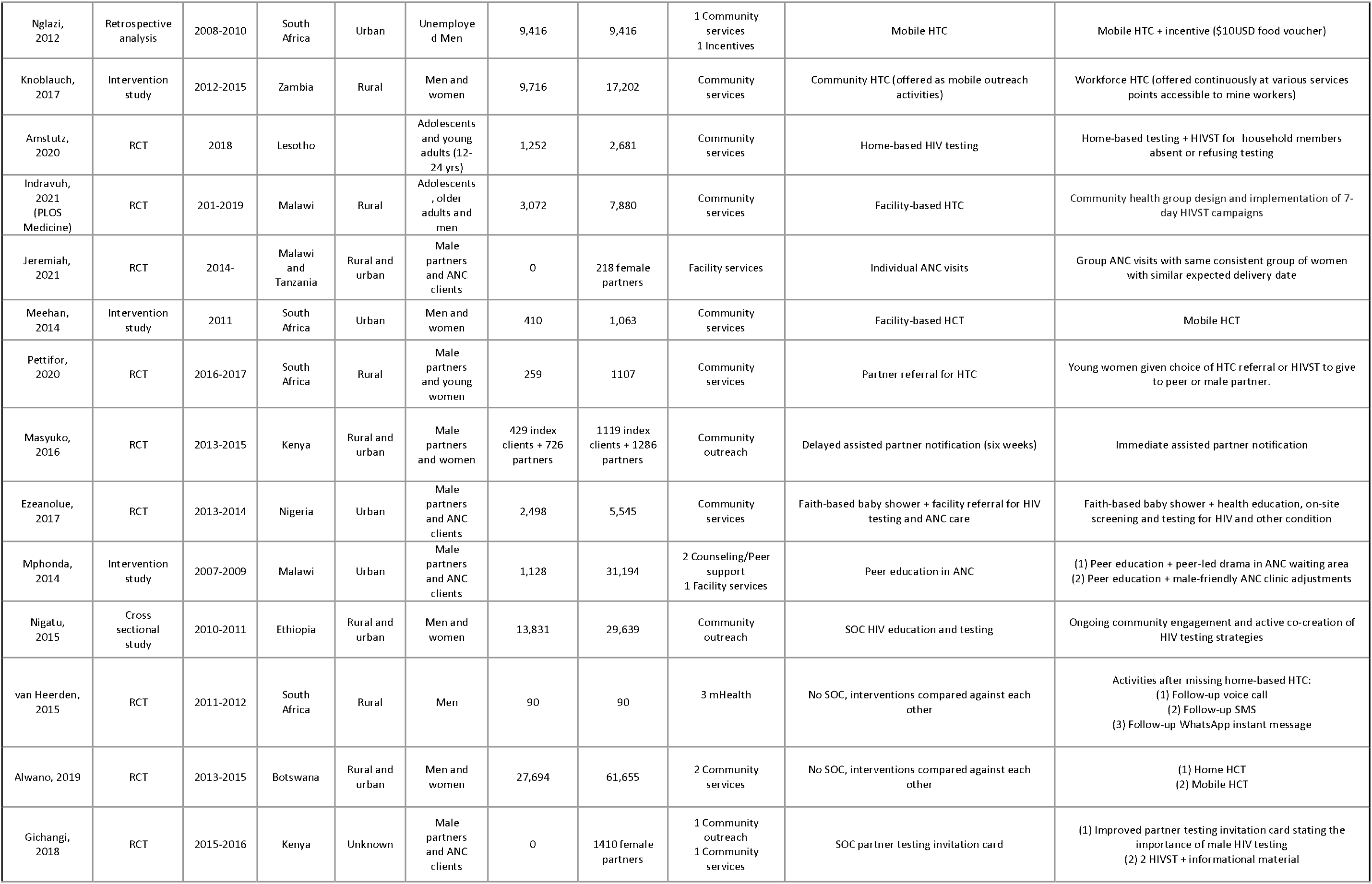
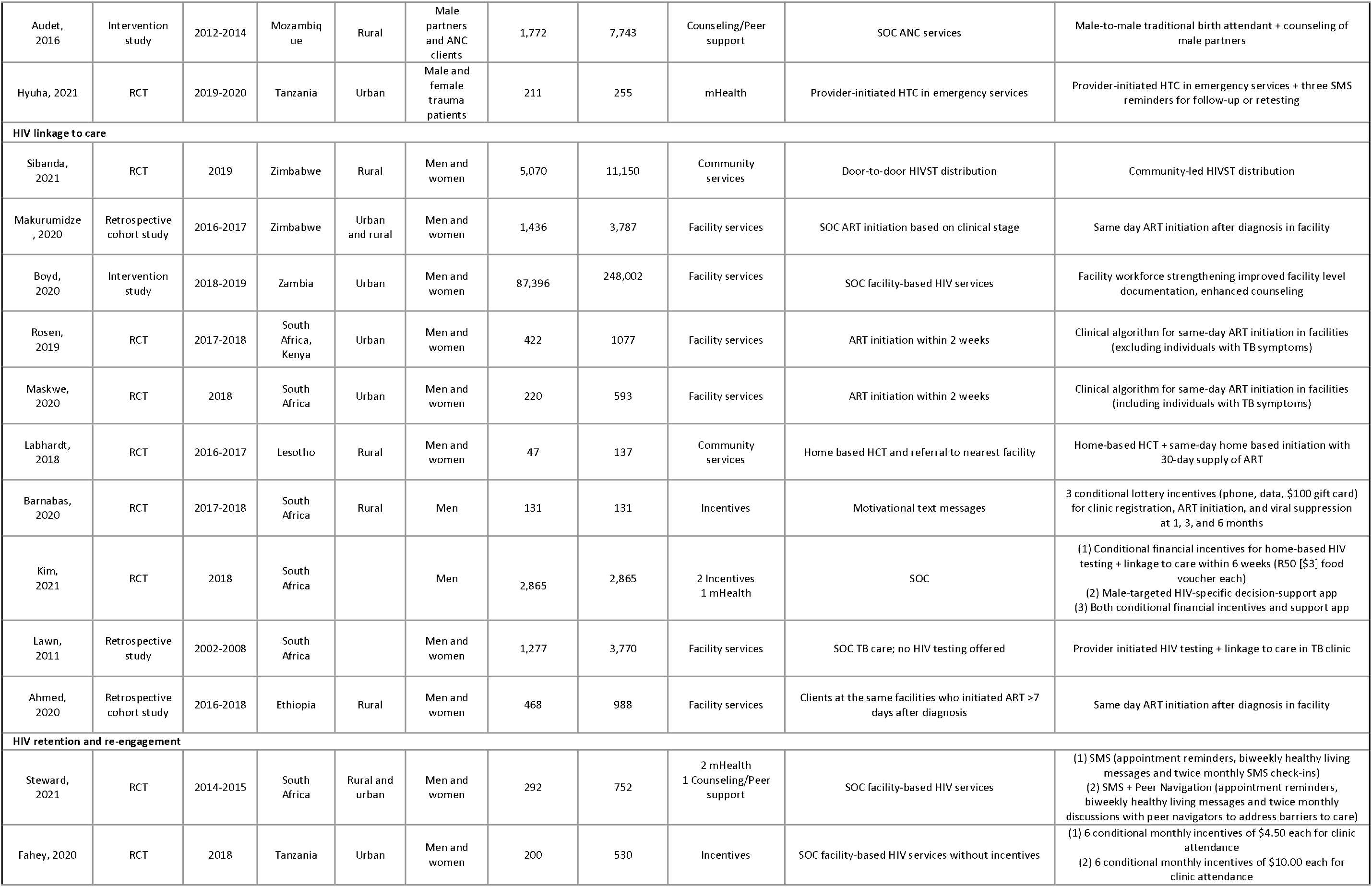
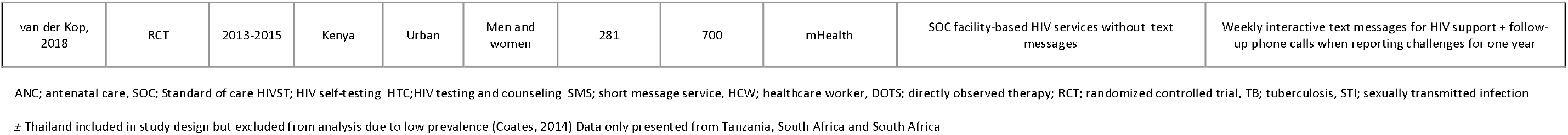
All studies, stratified by service domain.

**Table 6.**
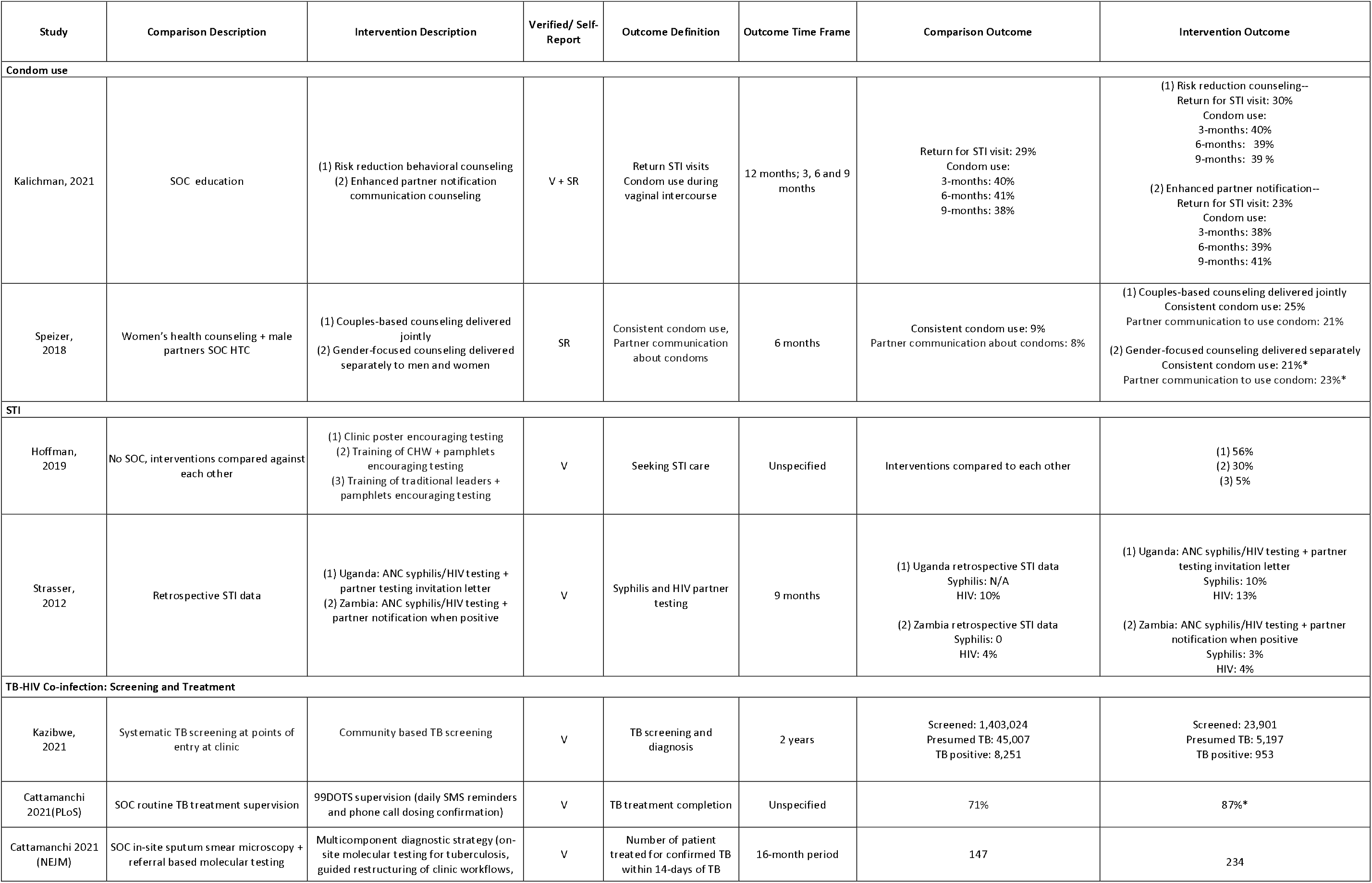
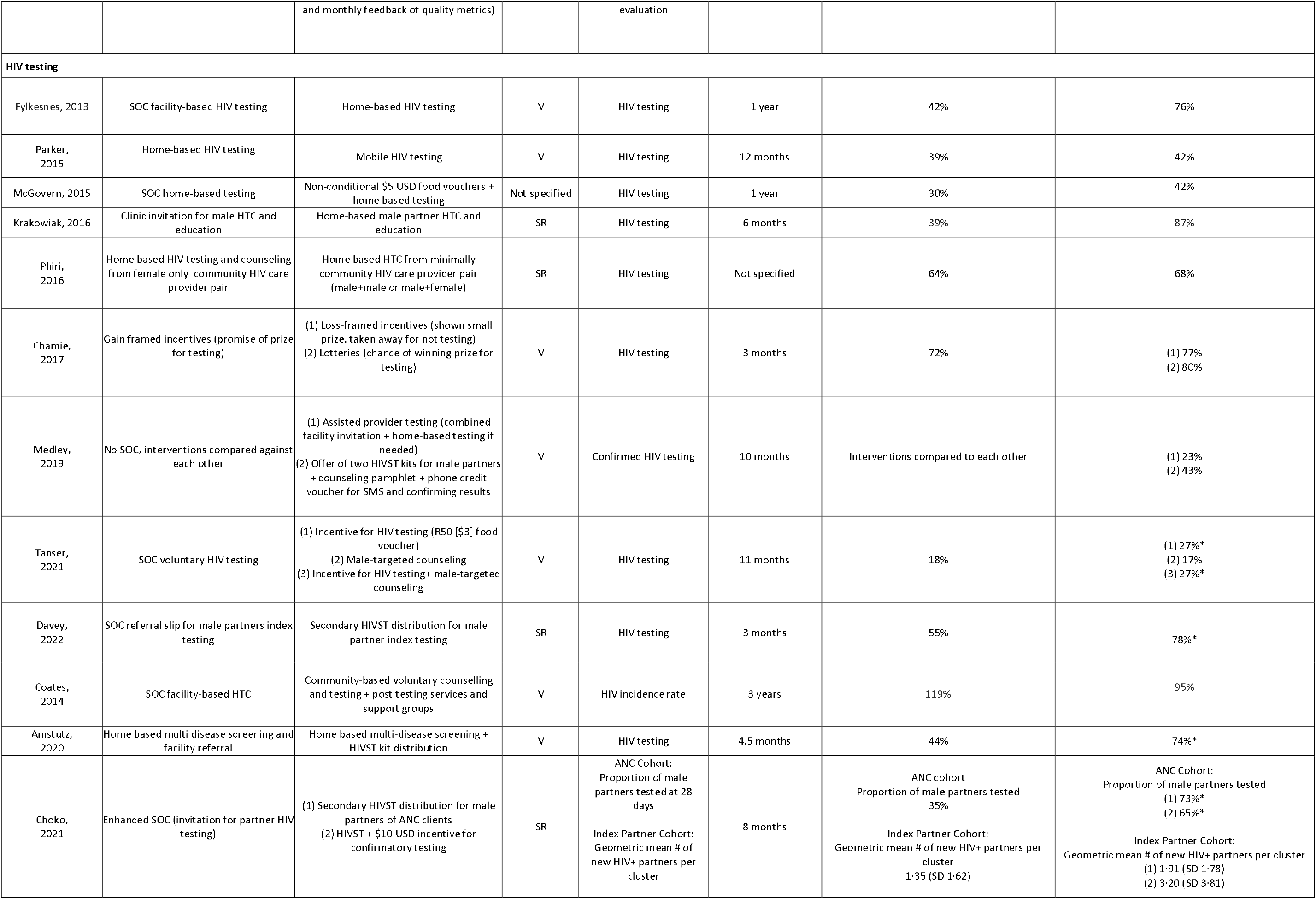
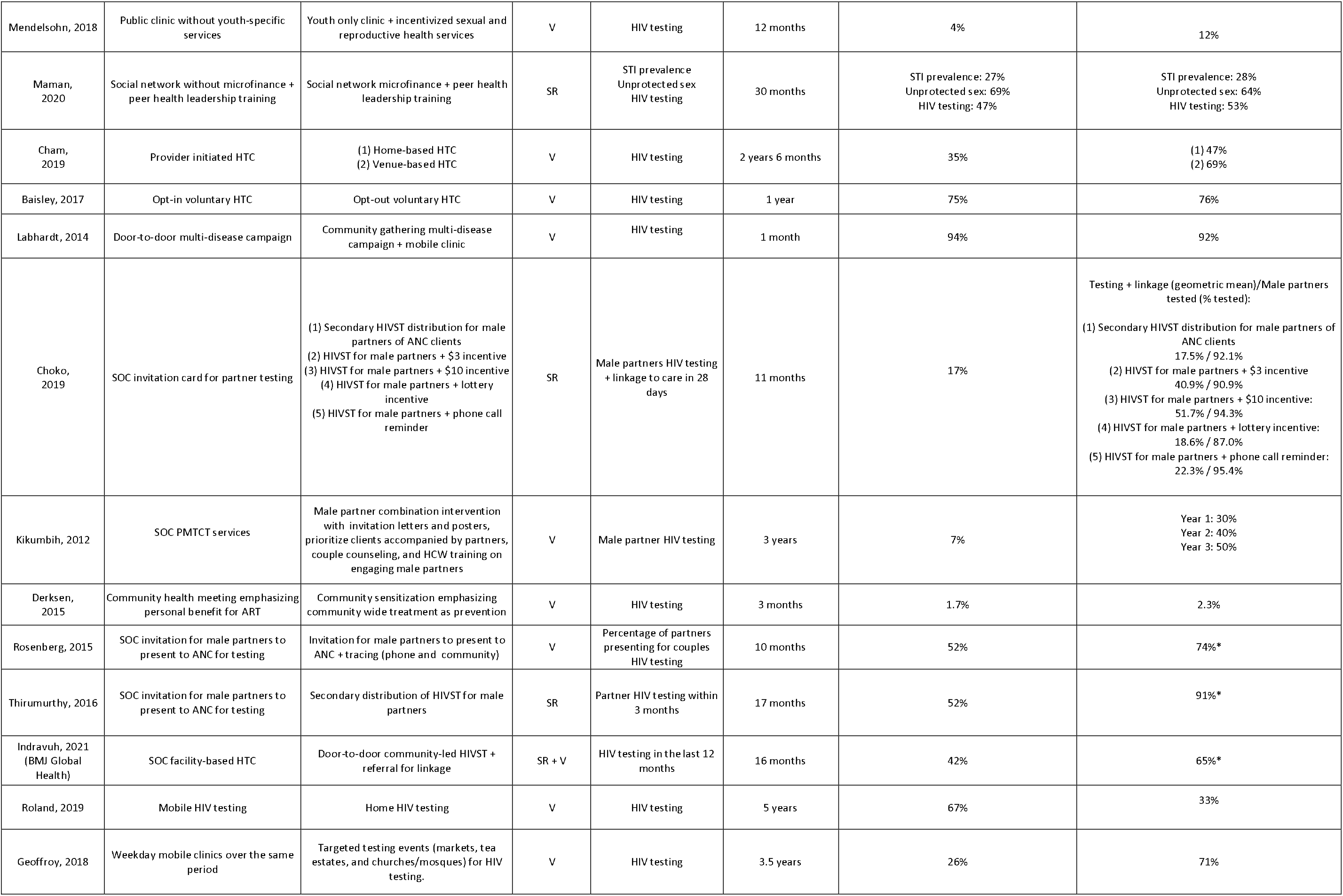
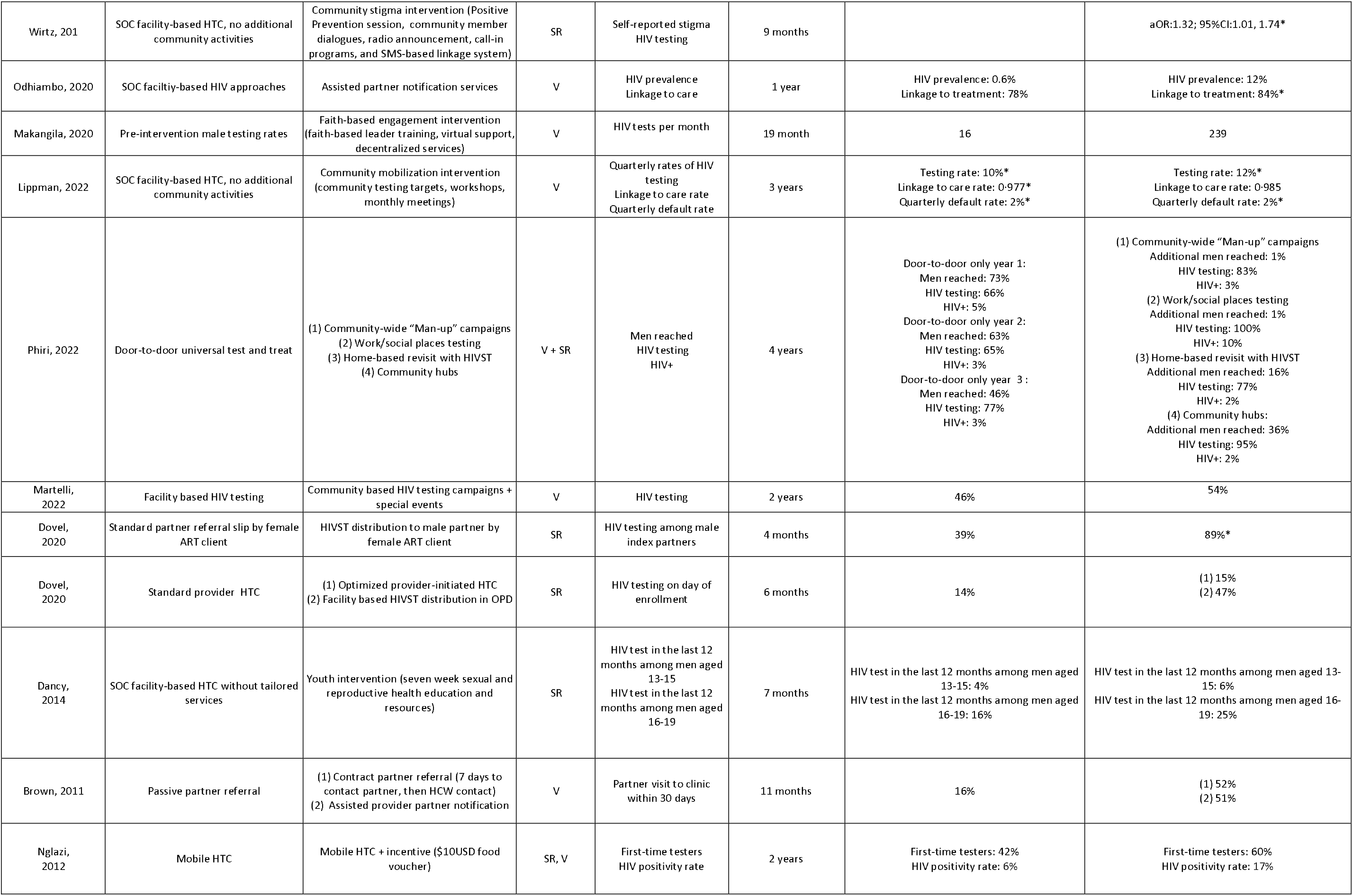
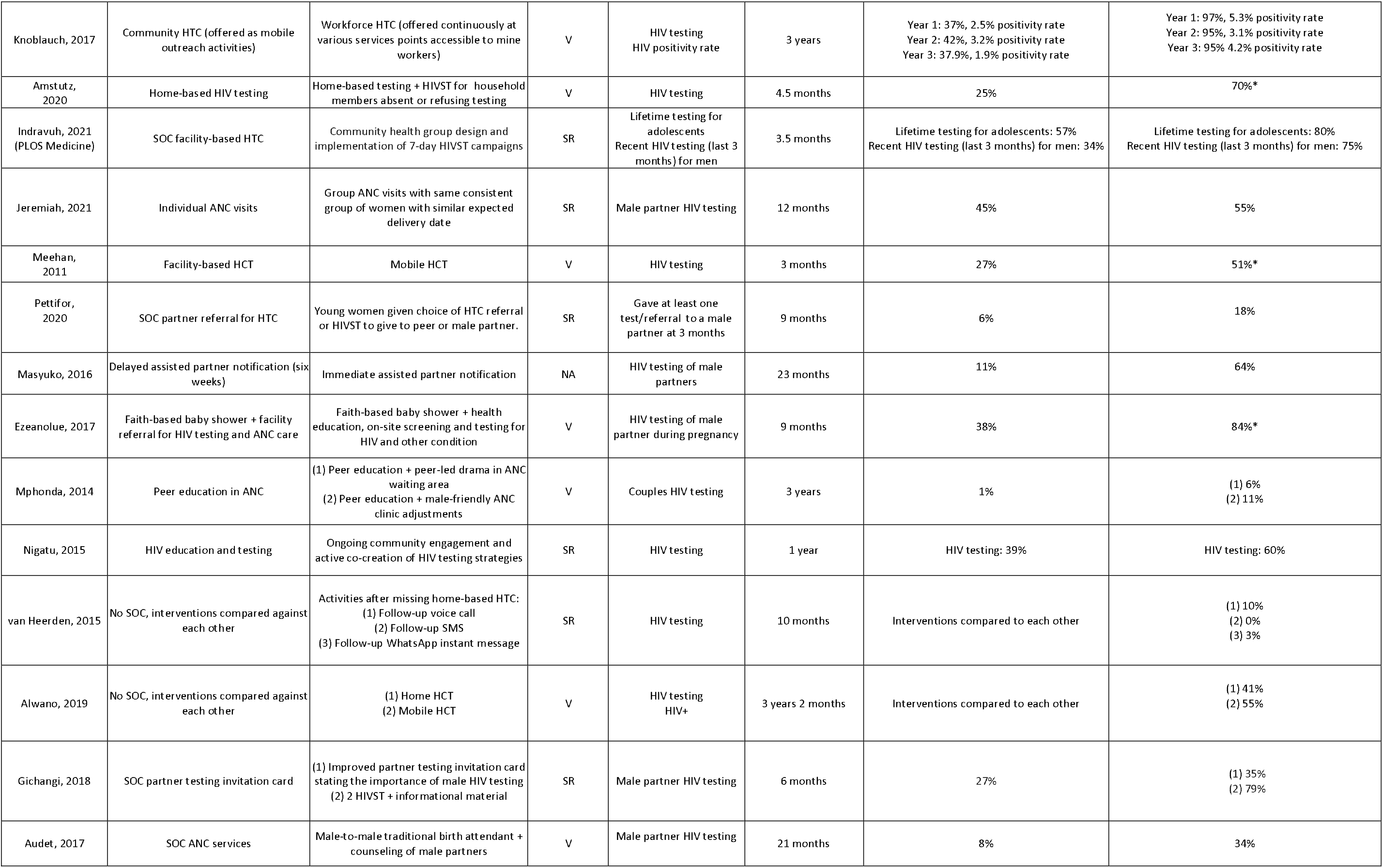
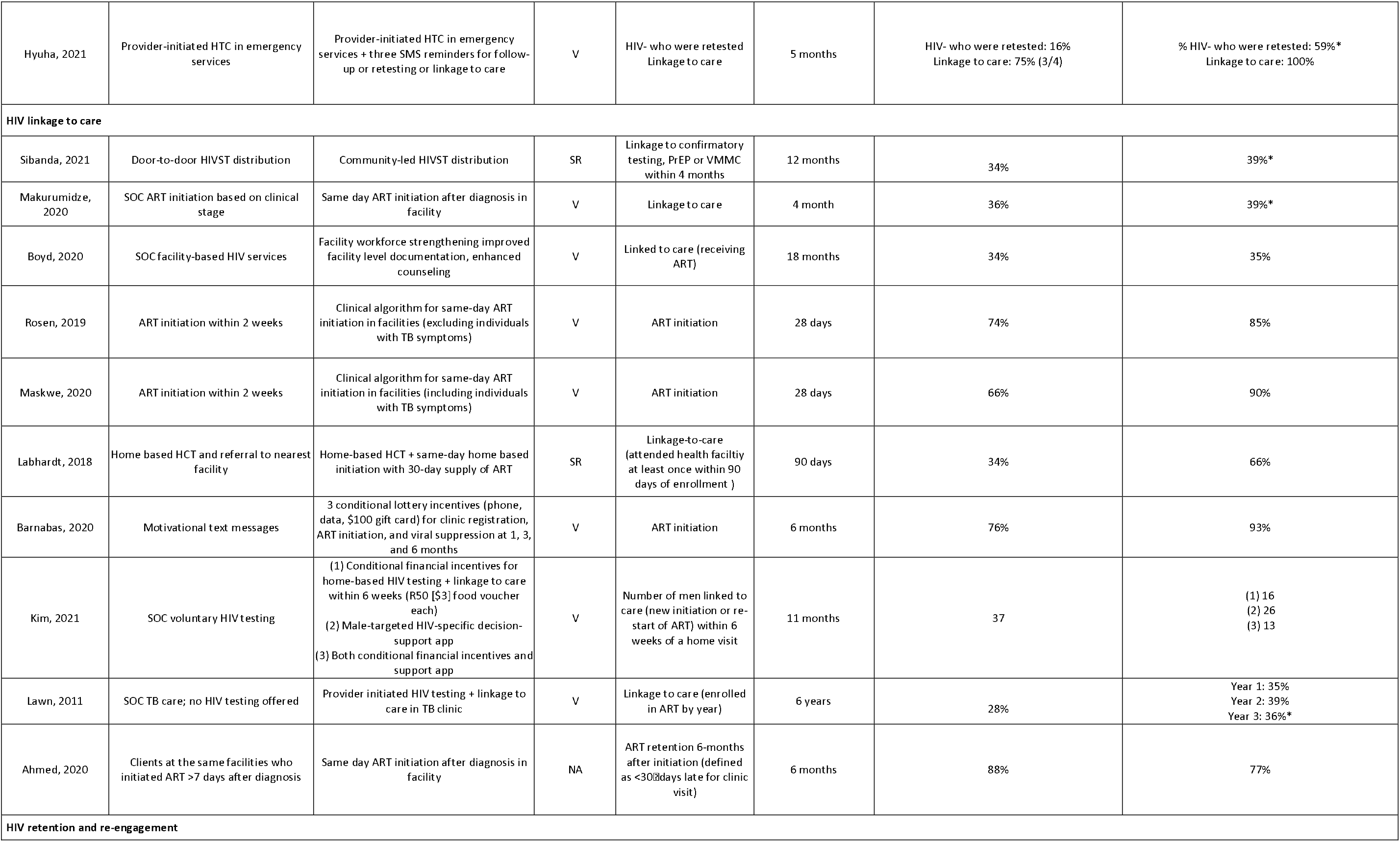
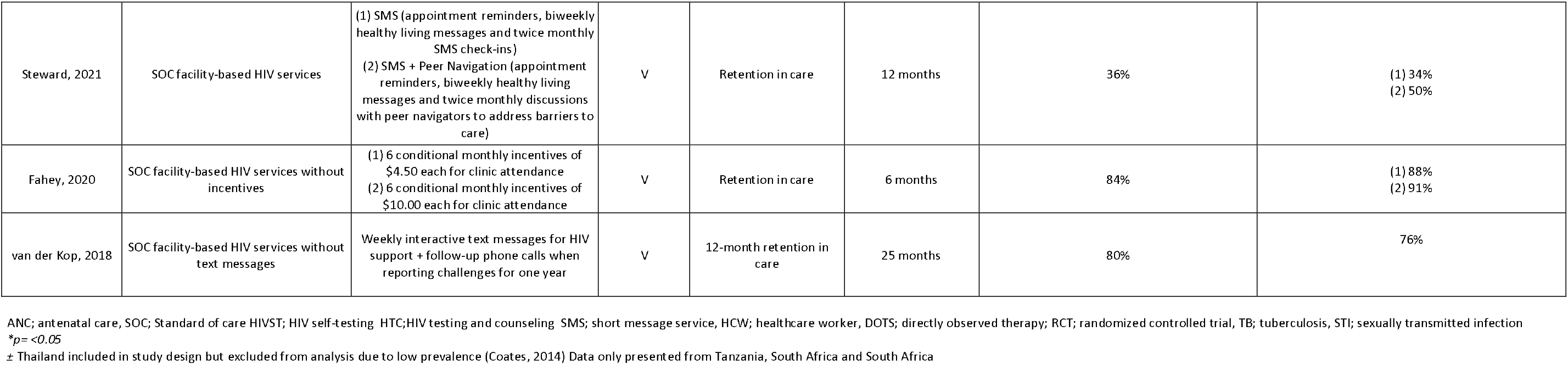
Study results, stratified by service domain.

### Facility Services

There were no interventions examining the impact of facility-based interventions on men’s use of condoms or PrEP services. Two interventions examined facility-based interventions for STI services and found that including clinic-based education (i.e., posters and pamphlets) and HCW training improved STI identification compared to traditional leader screening campaigns.^31^ One facility service intervention was included for TB-HIV co-infection and found that multicomponent diagnostic strategies improved co-infection diagnostics.^32^

Seven facility-based interventions focused on HIV testing as the primary outcome.^33–38^ Youth clinics,^33^ HCW sensitization on engaging male partners,^35^ and male-friendly spaces within ANC clinics^37,38^ improved men’s testing uptake. Offering opt-out HIV testing to men already attending health facilities has a wide reach but minimal impact^34,36^ unless paired with HIV self-testing (HIVST) strategies, which can dramatically increase testing coverage for men attending facilities.^36^

Six facility-based interventions focused on linkage to ART services after a reactive HIV test.^39–44^ Same-day initiation is strongly associated with linkage,^39,41,42^ although there is mixed evidence on impact on retention.^44^ Interventions that improve facility documentation to support enhanced counseling increased ART initiation,^40^ as did integrating TB and HIV services within the TB clinic.^43^

There were no facility-based interventions focused on improving retention.

### Community Services

There were no interventions examining the impact of community service interventions on use of condoms, PrEP, or STI services. One intervention for TB-HIV co-infection found that community-based TB screening increased screening efforts but decreased presumed TB and TB positive cases identified.^45^

There were a total of 39 community service interventions focused on HIV testing – 17 interventions distributed HIVST to men (primary or secondary distribution),^46–58^ 15 provided community/workforce testing (non-HIVST),^52,53,55,59–71^ and 11 provided home-based HIV testing services.^46,54,61,69,72–75^ Community services outperformed standard of care facility services^48,52,60,61,65,67,72,73^ and mobile and targeted venue testing such as work were more cost efficient and had greater reach than home-based testing.^53,59,61,63,69,70,74^ Notably no study compared a community service to a facility-based intervention (outside standard of care). Faith-based events and events around pregnancy can increase men’s testing during critical life periods.^63,64,68^

Primary distribution of HIVST increased reach testing.^48,52,53,55^ Secondary HIVST distribution increased testing among men when given to female partners living with HIV as part of index testing,^47,58^ ANC clients,^49–51,57^ young women,^56^ and when HIVST was left for male adults and adolescents who were not home during door-to-door distribution.^48,54^

Only one community service intervention examined the impact on linkage for men. Same-day initiation at home immediately after home-based testing increased ART initiation rates.^62^ No community service interventions examined the impact on men’s retention in HIV care.

### Community Outreach

There were no interventions examining the impact of community outreach interventions on use of condoms, PrEP or TB-HIV co-infection. Three community outreach interventions for STI testing or treatment were found.^31,76^ Community sensitization through traditional leaders was not as effective as facility-based sensitization for increasing men’s treatment for STI services,^31^ while partner invitation letters for STI + HIV testing of male partners of ANC clients (using an index testing approach) showed little to no impact on syphilis testing.^76^

There were 16 interventions for HIV testing with community outreach.^35,53,57,64,71,77–84^ Most were male partner testing strategies and reached men through invitation letters and a variety of community sensitization strategies.^35,57,78,80,82,83^ Partner invitations combined with male-friendly services at the health facility or active tracing were usually effective.^35,78,82,83^ One study found that simply distributing HIVST kits dramatically increased men’s testing compared to partner invitations.^57^ A few interventions used community sensitization to improve knowledge and motivation for HIV services,^53,64,71,84^ such as expanding knowledge of treatment as prevention,^77^ reducing stigma^79^ and youth-specific education,^81^ with mixed impact.

There were no interventions examining community outreach interventions for ART initiation and retention.

### Counseling / Peer Support

Four interventions examined the impact of counseling and/or peer support on condom use – all found that couples and male-only counseling groups increased condom use and partner communication about condoms.^85,86^ Three interventions examined the impact of counseling and/or peer support on HIV testing. Peer education and mentorship increased couples testing for male partners of ANC clients.^38,87^ Peer navigation and mentorship also improved ART retention when compared to standard of care and SMS messages alone.^88^

### Incentives

Twelve interventions examined the impact of incentives on HIV testing, with mixed findings.^46,49,50,66,89–92^ Incentives improved HIVST result reporting.^46,49,50^ Three interventions examined incentives for ART linkage with mixed results.^93,94^ One intervention examined the impact of incentives on retention.^95^

### mHealth

One mHealth intervention examined the impact on TB treatment completion among TB/HIV co-infections and found that SMS reminders and phone calls for direct observation therapy improved treatment completion when implemented with fidelity.^96^ mHealth alone showed mixed effects on HIV testing^91,97^ but mHealth in combination with other interventions were effective.^79,91^

## Discussion

In this scoping review of interventions, we were unable to identify cross-cutting strategies to reach men across HIV and related health services in sub-Saharan Africa, largely because there is little evidence outside HIV testing interventions. Nearly half of all interventions targeted HIV testing, 13% ART initiation, and 6% retention, while only a handful targeted condom use, STIs or TB-HIV co-infection. Over a quarter of interventions targeted male partners, usually of ANC clients, and only 7 interventions exclusively targeted men as clients. Almost half of interventions took place in the community, with about one-fifth focusing on changes to facility services and another one-fifth providing community outreach. Few interventions focused on counseling and peer support or mHealth strategies. The limited evidence available points to the fact that men need convenience, active outreach, and improved experiences with health services. The same principles may apply to all services intended to reach men, including sexual health and TB-HIV coinfection services.

There is extensive knowledge about community-based services, especially for HIV testing. Self-testing strategies that are convenient and give men autonomy are highly effective and may provide important lessons for other services. For example, STI and TB services may promote self-care through STI self-testing ^98^ and self-monitoring TB adherence. HIVST may also be used to promote PrEP uptake,^99^ although there is no evidence to date for men..

Sex disaggregation within manuscripts and conference abstracts is still not commonplace. The lack of sex-disaggregated outcomes is especially challenging given that men in SSA are often underrepresented in HIV studies. Multiple studies in our scoping review found that men’s outcomes were much worse than women, even in intervention arms. We strongly recommend that sex-disaggregated results become part of all primary data analysis plans and be presented whenever possible, including primary outcome articles and abstracts.

Evidence on interventions for men in SSA have several glaring gaps that must be addressed. First, there is very little evidence about interventions to improve linkage to care, retention, condom use, PrEP, STIs, and TB-HIV coinfection among men. There is almost no synergy across services, which may partly explain why so little progress has been made to improve men’s use of services to date. We know from qualitative and description studies what men need to make health services work for them. Easy access, quality services, positive patient-provider interactions, and for some men, tailored direct client support are required.^15,100,101^ However, we know very little about how to meet these needs for men outside of HIV testing. HIV testing services are the starting point for evidence since that’s where most evidence has been collected. But it should be the springboard for producing evidence on cross-service, holistic approaches to engaging men. We have more work to do to understand how to make cross-cutting improvements across services. Multi-disease approaches for HIV and NCDs have proven effective within community screening interventions^102^ and should be explored, particularly for HIV and related services such as prevention, STI diagnosis and treatment and TB-HIV co-infection.

Second, there is very little evidence on counseling and peer support for men, and nearly no evidence on how to improve quality services, especially HCW-client interactions, client agency, and client satisfaction with services received. Emerging evidence shows that equitable, positive, and trusting HCW interactions are critical to men’s continued use of services, as well as men’s satisfaction with services.^100,103,104^ Recent studies suggest that positive interactions with peer HCWs and HIV messaging that resonates with men’s priorities can lead to positive ART retention outcomes^105,106^ and should be explored for other health services. Further research is needed to understand how to best support positive interactions with HCWs and equip HCWs to interact with men well. Across the region, HCWs report implicit bias toward men as clients, misunderstand men and are unsure how to engage men well. ^107–109^ HCWs often describe men as “difficult”, “too powerful” and simply unable to be “good clients.”^107^ Misconceptions and implicit bias stem from a lack of understanding, experience, and professional skills/tools to engage a client population well. This can change. One strategy is to provide HCW sensitization trainings and job aids on how to interact with men as clients.

Our scoping review has several limitations. First, nearly all evidence from southern and eastern Africa and may not be representative of what works in western and northern Africa. Second, we exclude articles and abstracts without a comparison group to ensure quality. Numerous programmatic and qualitative evaluations are now being completed and may provide useful insights into the needs of men and strategies that work – this literature should be explored further regarding cross-cutting strategies to reach men. Third, we include articles across a wide range of years. Effective strategies may shift over time as local epidemics change. Finally, we only included articles that were published in English.

## Conclusion

This review highlights the need for additional research on cross-cutting strategies to improve men’s engagement in HIV and related health services. The limited evidence available suggests that convenient services, actively engaging men, and providing positive experiences with health services largely improve service utilization. Additional evidence is needed for PrEP use and non-HIV services (such as STI and TB co-infection).

## Data Availability

All data presented in this scoping review are publicly available from their original sources.

## Acknowledgements

We are to grateful to all members of the WHO and UNAIDS Men and HIV technical Working Group (MENHT) who supported this work and to Misheck Mphande and Isabella Robson who supported collection of articles.

